# Maternal and Child Health Trends in Nigeria: A Scoping Review of NDHS 2018 vs. NDHS 2023

**DOI:** 10.1101/2025.05.18.25327864

**Authors:** Mordecai Oweibia, Christopher Ononiwu Elemuwa, Tarimobowei Egberipou, Gift Cornelius Timighe, Peresuodei Sylvanus, Tuebi Richard Wilson

## Abstract

**Background:** Despite decades of intervention, Nigeria continues to face critical challenges in maternal and child health, ranking among the highest globally for maternal mortality and under-five deaths. Sustainable Development Goals (SDGs) 3.1 and 3.2 call for substantial reductions in these outcomes by 2030. Assessing national progress requires not only measuring quantitative indicators but also understanding contextual and systemic barriers that shape health access and equity.

**Methods:** This study adopted a mixed-methods design combining descriptive quantitative analysis of the NDHS 2018 and 2023–24 datasets with a thematic qualitative analysis using NVivo software. Five focus areas were assessed: maternal and under-five mortality, immunization coverage, exclusive breastfeeding and malnutrition, antenatal care and skilled birth attendance, and treatment-seeking behavior for childhood illnesses. Quantitative results were complemented with thematic insights drawn from NDHS narrative and policy sections to capture structural and community-level determinants.

**Results:** The under-five mortality rate declined from 132 to 102 deaths per 1,000 live births. Improvements were also observed in immunization coverage, exclusive breastfeeding (from 29% to 34%), and antenatal care attendance (from 57% to 68%). Skilled birth attendance rose to 52%, and treatment-seeking for childhood illnesses increased. However, maternal mortality data for 2023 were unavailable, and persistent disparities, particularly in rural and northern regions, remain evident. Qualitative analysis revealed themes of access barriers, mistrust in services, inconsistent program implementation, and gaps in data systems.

**Conclusion:** Nigeria has made measurable but uneven progress in maternal and child health. Structural challenges, regional inequities, and weak data governance continue to hinder national gains. A combination of integrated service delivery, targeted social protection, and equitable policy implementation is needed to accelerate progress toward SDG 3 targets.

## 1.0 INTRODUCTION

### 1.1 Background of the Study

Maternal and child health remains a fundamental cornerstone of sustainable development and public health in any nation, especially in developing countries like Nigeria, where morbidity and mortality rates remain unacceptably high (Elemuwa *et al.,* 2024). Despite substantial national efforts, global partnerships, and the existence of strategic frameworks, the health outcomes for mothers and children in Nigeria have been largely unsatisfactory and unevenly distributed across the country’s six geopolitical zones (FMoHSW, NPC, & ICF, 2024). These disparities reflect deep-rooted systemic challenges, including inequitable access to health services, socio-cultural barriers, poor funding, and inadequate infrastructure (Okeke *et al.,* 2023).

The Sustainable Development Goals (SDGs), established in 2015 by the United Nations, represent a universal call to action to end poverty, protect the planet, and ensure that all people enjoy peace and prosperity (Mordecai, 2023). Specifically, SDG 3.1 aims to reduce the global maternal mortality ratio (MMR) to less than 70 per 100,000 live births, while SDG 3.2 focuses on ending preventable deaths of newborns and children under five by 2030 (Mordecai *et al.,* 2023). Nigeria, as a signatory to the SDGs and a member of the United Nations, is expected to align its national development policies and health programming with these global targets (Akpan *et al.,* 2024).

According to the Nigeria Demographic and Health Survey (NDHS) 2018, the maternal mortality ratio was estimated at 512 deaths per 100,000 live births, one of the highest globally (NPC & ICF, 2019). Meanwhile, under-five mortality stood at 132 deaths per 1,000 live births, reflecting severe gaps in child survival interventions (FMoHSW, NPC, & ICF, 2024). The more recent NDHS 2023–24 Key Indicators Report shows that under-five mortality has declined to 102 per 1,000 live births, signaling modest improvements but still far above the SDG target of 25 per 1,000 (FMoHSW, NPC, & ICF, 2024).

One major determinant of these poor health outcomes is Nigeria’s fragmented healthcare delivery system. The federal, state, and local governments share overlapping responsibilities, often resulting in duplicated efforts, inefficient resource allocation, and disjointed service delivery (Mordecai *et al.,* 2023). These challenges are compounded by chronic underfunding of the health sector, with health expenditure consistently falling below the 15% benchmark stipulated in the 2001 Abuja Declaration (Mordecai *et al.,* 2024).

The issue of poverty remains one of the most significant contributors to poor maternal and child health outcomes in Nigeria. The 2022 National Multidimensional Poverty Index (MPI) revealed that over 63% of Nigerians, approximately 133 million people, live in multidimensional poverty, experiencing multiple deprivations in health, education, and living standards (Gyang *et al.,* 2023). This level of deprivation restricts access to healthcare services and nutritional resources essential for the health of women and children (Akpan *et al.,* 2024).

Another prominent barrier is the limited availability and accessibility of quality maternal and child health services. Many rural communities in Nigeria lack functional health centers, trained personnel, and essential drugs, leading to high rates of home deliveries, low immunization coverage, and delayed treatment for childhood illnesses (Umesi, 2023). In the 2023 NDHS report, only 52% of births were attended by skilled health personnel, a slight improvement from 43% in 2018, but still below optimal coverage (FMoHSW, NPC, & ICF, 2024).

The prevalence of communicable and preventable diseases continues to burden the pediatric population in Nigeria. According to the NDHS 2023–24, 23% of children under five experienced fever, and 9% had diarrhea in the two weeks preceding the survey, with treatment-seeking behavior improving slightly but still affected by geographical and financial constraints (FMoHSW, NPC, & ICF, 2024). Similarly, pneumonia and acute respiratory infections remain a concern, especially in malnourished children in the northern regions of the country (Elemuwa *et al.,* 2024).

Immunization coverage has seen modest improvements between 2018 and 2023. For example, DPT3 coverage rose from 50% to 57%, measles from 54% to 59%, and pentavalent vaccines from 50% to 56%, though still significantly below the World Health Organization’s recommended 90% coverage for herd immunity (FMoHSW, NPC, & ICF, 2024). Vaccine hesitancy, misinformation, and logistical failures continue to impede further progress (Mordecai *et al.,* 2023).

Malnutrition, a critical underlying factor in child mortality, remains alarmingly high. In 2023, stunting affected 32% of children under five, while 6% were wasted and 19% underweight only marginally better than 2018 figures (FMoHSW, NPC, & ICF, 2024). These figures are deeply concerning given the associated cognitive and physical developmental impairments that malnutrition causes (Kareem *et al.,* 2023).

On the maternal side, antenatal care (ANC) coverage has improved, with 68% of women receiving at least four ANC visits in 2023, compared to 57% in 2018 (FMoHSW, NPC, & ICF, 2024). However, ANC is often initiated late, with poor counseling and service quality further undermining its potential benefits (Mordecai, 2023). Inadequate access to emergency obstetric care, particularly cesarean sections and blood transfusions, continues to be a leading cause of preventable maternal deaths (Chidinma, 2024).

Cultural and religious beliefs significantly influence maternal health-seeking behavior in Nigeria. In several northern states, traditional norms restrict women’s mobility and decision-making power, requiring male permission before seeking health services (Elemuwa *et al.,* 2024). These sociocultural limitations are reinforced by low levels of female education and economic dependence, compounding the challenges of timely access to care (Mordecai, 2023).

Recognizing these challenges, the Nigerian government has introduced several national health strategies and interventions. These include the Integrated Maternal, Newborn, and Child Health (IMNCH) strategy, the Basic Health Care Provision Fund (BHCPF), and the National Strategic Health Development Plan II (NSHDP II) (Akpan *et al.,* 2024). These policies aim to increase the availability of health workers, strengthen PHCs, and reduce out-of-pocket expenditures for maternal and child health services (Agbonle *et al.,* 2022).

One of the most promising interventions in recent years has been the implementation of community-level structures such as the Ward Health System (WHS), which connects each health facility to a Health Facility Development Committee (HFDC) responsible for community mobilization and service oversight (Elemuwa *et al.,* 2024). This approach has proven effective in enhancing accountability and increasing community ownership of health programs, particularly immunization campaigns and COVID-19 responses (Kitila *et al.,* 2022).

The COVID-19 pandemic, while disrupting routine health services, offered a stress test for Nigeria’s health system. The accelerated rollout of vaccines showed that with political will, community engagement, and efficient logistics, even fragile systems can deliver high-impact interventions (Babatunde *et al.,* 2024). Lessons from the COVID-19 response are now being used to design more resilient maternal and child health delivery systems (Mordercai *et al.,* 2023).

Access to reliable data remains a cornerstone of effective health planning and monitoring. The NDHS, conducted every five years, remains Nigeria’s most authoritative source of health and demographic data (NPC & ICF, 2019; FMoHSW, NPC, & ICF, 2024). However, the lack of disaggregated data in preliminary reports and delays in full survey releases pose significant barriers to timely policymaking (Mordecai, 2023).

The cumulative evidence presented by the NDHS and academic literature highlights the interdependency of maternal and child health indicators. Poor maternal health invariably translates into poor child outcomes, as seen in the transmission of HIV, poor birth spacing, low birth weight, and neonatal complications (Babajide *et al.,* 2023). Therefore, improving maternal health is not only a moral imperative but also an investment in child survival and national development (Akpan *et al.,* 2024).

### 1.2 Statement of the Problem

Despite decades of health sector reform and strategic planning, Nigeria continues to face some of the highest maternal and child mortality rates globally, with uneven progress across its regions. As of the 2018 NDHS, the maternal mortality ratio stood at 512 deaths per 100,000 live births, far exceeding the SDG 3.1 target of fewer than 70 deaths per 100,000 (NPC & ICF, 2019). Though under-five mortality declined from 132 to 102 per 1,000 live births between 2018 and 2023, this figure remains unacceptably high by global standards and is indicative of slow progress toward SDG 3.2 (FMoHSW, NPC, & ICF, 2024). Disparities persist between urban and rural populations, as well as among Nigeria’s geopolitical zones, with northern states continuing to bear a disproportionate burden of child deaths due to malnutrition, limited access to immunization, and low coverage of skilled birth attendance (Akintunde *et al.,* 2023). Access to maternal services is often impeded by factors such as poverty, sociocultural norms, poor road infrastructure, and lack of emergency transportation, which collectively delay or prevent care-seeking behavior, particularly in remote communities (Jeremiah *et al.,* 2023).

Further compounding the issue is Nigeria’s fragmented health system, characterized by weak intergovernmental coordination, underfunded primary healthcare centers, and a chronic shortage of trained health personnel in underserved areas (Gyang *et al.,* 2023). Although skilled birth attendance and antenatal care coverage have improved slightly between 2018 and 2023, these gains remain insufficient and poorly distributed across states (FMoHSW, NPC, & ICF, 2024). Health system resilience remains low, with frequent supply stock outs, infrastructure decay, and gaps in maternal emergency preparedness, such as blood banking and surgical capacity (Babatunde *et al.,* 2023).

Additionally, the unavailability of updated national-level maternal mortality data in the preliminary 2023 NDHS report limits the ability to evaluate progress comprehensively (Ajayi *et al*., 2023). Without targeted, data-informed interventions that address both systemic and contextual barriers, Nigeria is unlikely to meet its national and global targets for reducing maternal and child mortality (Mordercai *et al.,* 2024).

### 1.3 Aim and Objectives of the Study

The aim of this study is to assess the trends in maternal and child health outcomes in Nigeria by comparing indicators from the 2018 and 2023–24 Nigeria Demographic and Health Surveys (NDHS), in alignment with Sustainable Development Goals 3.1 and 3.2. The study seeks to determine the extent of progress made in improving health service coverage, reducing mortality, and addressing malnutrition and health-seeking behaviors in children under five and their mothers.

**Specific Objectives:**

**1.** To compare maternal mortality ratios (MMR) and under-five mortality rates (U5MR) between the 2018 and 2023 NDHS to evaluate progress toward SDG 3.1 and SDG 3.2.
**2.** To analyze trends in neonatal and child immunization coverage (DPT3, measles, pentavalent vaccines) between 2018 and 2023.
**3.** To examine changes in exclusive breastfeeding rates and malnutrition indicators (stunting, wasting, underweight) among children under five.
**4.** To assess antenatal care (ANC) and skilled birth attendance (SBA) coverage and their correlation with neonatal health outcomes, including early neonatal mortality.
**5.** To evaluate the prevalence of common childhood illnesses (malaria, diarrhea, pneumonia) and associated treatment-seeking behavior during the 2018–2023 period.

### 1.4 Research Questions

To guide the study, the following research questions are posed:

**1.** What are the comparative trends in maternal mortality and under-five mortality in Nigeria between 2018 and 2023?
**2.** How has immunization coverage for major childhood vaccines changed over the five-year period?
**3.** What are the observed shifts in exclusive breastfeeding practices and malnutrition indicators among children under five?
**4.** What relationship exists between ANC/SBA coverage and neonatal survival outcomes in the two NDHS reporting years?
**5.** Have treatment-seeking behaviors for childhood illnesses improved, and what does the data suggest about healthcare access and utilization?

### 1.5 Significance of the Study

This study provides a timely and evidence-based analysis of Nigeria’s progress in improving maternal and child health outcomes using two key national datasets: NDHS 2018 and 2023–24. By examining trends across critical indicators such as maternal and under-five mortality, immunization, nutrition, and healthcare access it offers actionable insights for policymakers, program designers, and public health practitioners. The findings will support the development of targeted interventions to address persisting disparities across regions and socio-economic groups, particularly in achieving SDG 3.1 and 3.2. Academically, the study contributes to the literature on health systems performance in low-resource settings by providing a comparative, data-driven approach to assessing health outcomes over time. It also informs future monitoring and evaluation frameworks by highlighting gaps in service delivery and policy implementation. Ultimately, the research serves as a tool for improving equity, efficiency, and accountability in maternal and child health governance in Nigeria.

### 1.6 Definition of Terms

- Maternal Mortality Ratio (MMR): The number of maternal deaths per 100,000 live births during a specified time period, typically due to complications from pregnancy, childbirth, or postpartum causes.
- Under-Five Mortality Rate (U5MR): The probability (per 1,000 live births) that a child will die before reaching the age of five, often used as a measure of child health and health system performance.
- Nigeria Demographic and Health Survey (NDHS): A nationally representative survey conducted periodically to provide reliable data on population health indicators, fertility, family planning, maternal and child health, and service coverage.
- Antenatal Care (ANC): Routine health check-ups and services provided to pregnant women to monitor the progress of pregnancy and identify complications early.
- Skilled Birth Attendance (SBA): Delivery of a baby with the assistance of a trained health professional such as a doctor, nurse, or midwife, capable of handling normal deliveries and recognizing complications.
- Exclusive Breastfeeding: The practice of feeding an infant only breast milk (no other liquids or solids) for the first six months of life, as recommended by WHO and UNICEF.
- Immunization Coverage: The proportion of children who receive vaccinations against preventable diseases, typically measured by age-appropriate completion of scheduled vaccines like DPT3, measles, and pentavalent.
- Malnutrition Indicators: Statistical measures of child nutritional status, including stunting (low height-for-age), wasting (low weight-for-height), and underweight (low weight-for-age).
- Treatment-Seeking Behavior: The actions taken by caregivers to seek medical care when children present symptoms of illness, including type and timeliness of health services accessed.

### 1.7 Limitations of the Study

While this study draws on the Nigeria Demographic and Health Survey (NDHS) data, which are nationally representative and widely validated, certain limitations are acknowledged. First, the NDHS 2023–24 data are based on the Key Indicators Report, which may not include detailed disaggregation by state or socioeconomic strata, limiting the depth of subnational analysis. Second, maternal mortality data from the 2023–24 survey were not available at the time of this review, making direct year-to-year comparison on that indicator infeasible. Third, the study is descriptive in nature and does not explore causal relationships. Finally, reliance on secondary data means that some contextual factors such as health facility quality or informal care use are not captured. These limitations, while significant, do not detract from the study’s ability to highlight important trends and inform future policy and research directions.

### 2.0 MATERIALS AND METHODS

This scoping review was conducted to map and compare maternal and child health trends in Nigeria using data from the 2018 and 2023–24 Nigeria Demographic and Health Surveys (NDHS). The methodology follows the framework of Arksey and O’Malley (2005), enhanced by Levac et al. (2010), and adheres to the PRISMA-ScR (Tricco *et al.,* 2018) and Joanna Briggs Institute (JBI) guidelines. The review was organized around five specific objectives covering mortality rates, immunization, malnutrition, maternal care, and childhood diseases.

### 2.1 Study Design

A scoping review methodology was chosen to provide a comprehensive overview of health indicator trends rather than assess the quality of intervention outcomes. This design enabled mapping of existing evidence, identifying progress and gaps, and informing policy recommendations. The approach involved the following steps:

- Identifying the research questions using the five stated objectives.
- Systematic selection and extraction of data from NDHS reports and related literature.
- Thematic mapping of trends and comparisons across two time points: 2018 and 2023.

### 2.2 Data Sources and Search Strategy

The primary data sources were:

- NDHS 2018 (NPC & ICF, 2019)
- NDHS 2023–24 Key Indicators Report (FMoHSW, NPC, & ICF, 2024)
- sources included:
- Peer-reviewed articles from databases such as PubMed, Scopus, Web of Science, and AJOL.
- Grey literature from WHO, UNICEF, Nigeria Federal Ministry of Health (FMoH), and the National Primary Health Care Development Agency (NPHCDA).

Search terms included combinations of:

- Maternal mortality, child health, NDHS Nigeria, exclusive breastfeeding, vaccination, malnutrition, skilled birth, under-five mortality, SDG 3.1, and SDG 3.2.

Boolean operators ("AND", "OR") were used to filter relevant records. All searches were restricted to publications between 2015 and 2025, written in English.

### 2.3 Eligibility Criteria

- Inclusion Criteria:

- Studies or datasets focusing on Nigeria between 2018 and 2023.
- Quantitative data on MMR, U5MR, immunization, malnutrition, ANC/SBA, or child illnesses.
- Qualitative data highlighting barriers to care, policy shifts, or program implementation.
- Exclusion Criteria:

- Studies unrelated to maternal or child health.
- Non-English documents.
- Reports outside the specified timeframe or lacking national-level relevance.

### 2.4 Data Extraction

Data were extracted manually using a structured Microsoft Excel template, organized by objective and thematic category (Peters *et al.,* 2015). Extracted variables included:

- Maternal health: MMR, ANC coverage (4+ visits), skilled birth attendance, postnatal care.
- Child health: U5MR, neonatal mortality, immunization (DPT3, measles, pentavalent), breastfeeding rates.
- Nutrition: Rates of stunting, wasting, and underweight in children under five.
- Childhood illness: Prevalence of malaria, diarrhea, pneumonia, and care-seeking behavior.
- Contextual data: Socioeconomic disparities, rural–urban differences, geopolitical zone breakdowns, and reported policy efforts.

NVivo software was used to thematically code any qualitative insights, particularly for contextual barriers or enablers described in policy or narrative sections of the NDHS reports.

### 2.5 Data Analysis

**a.** Quantitative Analysis

A descriptive comparison of indicators was conducted for 2018 and 2023 using:

- Percentages and frequency tables
- Cross-tabulations by zone, state, and urban–rural residence

Where applicable, change over time was calculated using the percentage change formula:

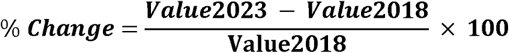

Graphical representations, including line graphs, bar charts, and heat maps were generated using Microsoft Excel. Data visualization focused on highlighting trends, gaps, and regional disparities.

**b.** Qualitative Analysis

Narrative insights were coded using thematic analysis to capture recurring themes such as:

- Cultural barriers to ANC utilization
- Policy reforms on routine immunization
- Equity and accessibility issues in rural and conflict-prone areas

**c.** Comparative Benchmarking

- Nigerian data were benchmarked against WHO/UNICEF targets (e.g., ≥90% immunization coverage, ANC ≥4 visits).
- Trends were also compared with regional peers (e.g., Ghana, Kenya) where relevant secondary data existed.

### 2.6 Quality Assessment

Quality of extracted data was assessed using the Joanna Briggs Institute (JBI) appraisal tools. NDHS datasets were recognized as gold standard population-based surveys, though limitations such as recall bias, non-sampling errors, and data gaps in insecure regions were acknowledged and accounted for in the interpretation.

### 2.7 Ethical Considerations

The review did not involve primary data collection or interaction with human participants. All data were obtained from publicly accessible documents and were cited accordingly. As such, ethical approval was not required.

**Mathematical Expressions and Symbols**

The following formulas were used for the quantitative analysis in this review:

**1.** Percentage Change Formula

To calculate change in health indicators between 2018 and 2023:

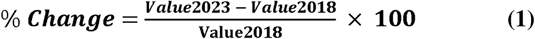

**2.** Trend Slope (if regression is applied in deeper analysis) (Wooldridge, 2015):

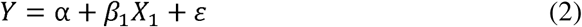

Where:

- Y = Health outcome (e.g., immunization rate)
- X_1_ = Time (survey year)
- α = Intercept
- β_1_ = Coefficient of change
- ε = Error term

Note: Formula 2 will only be applied if further inferential analysis is conducted beyond the descriptive level.

### 3.0 RESULTS

### 3.1 MMR and U5MR Comparison (2018 vs 2023)

The under-five mortality rate (U5MR) in Nigeria showed a notable decline from 132 deaths per 1,000 live births in 2018 to 102 deaths per 1,000 live births in 2023, as reported in the NDHS surveys. This represents an approximate 22.7% reduction over the five-year period, indicating measurable progress toward Sustainable Development Goal 3.2 (ending preventable deaths of children under five) (NPC & ICF, 2019; FMoHSW, NPC, & ICF, 2024).

However, the maternal mortality ratio (MMR), which was reported at 512 deaths per 100,000 live births in 2018, was not included in the 2023–24 NDHS Key Indicators Report. This omission limits direct year-to-year comparison for SDG 3.1 (reducing maternal deaths to fewer than 70 per 100,000 live births by 2030).

**Table 1:**
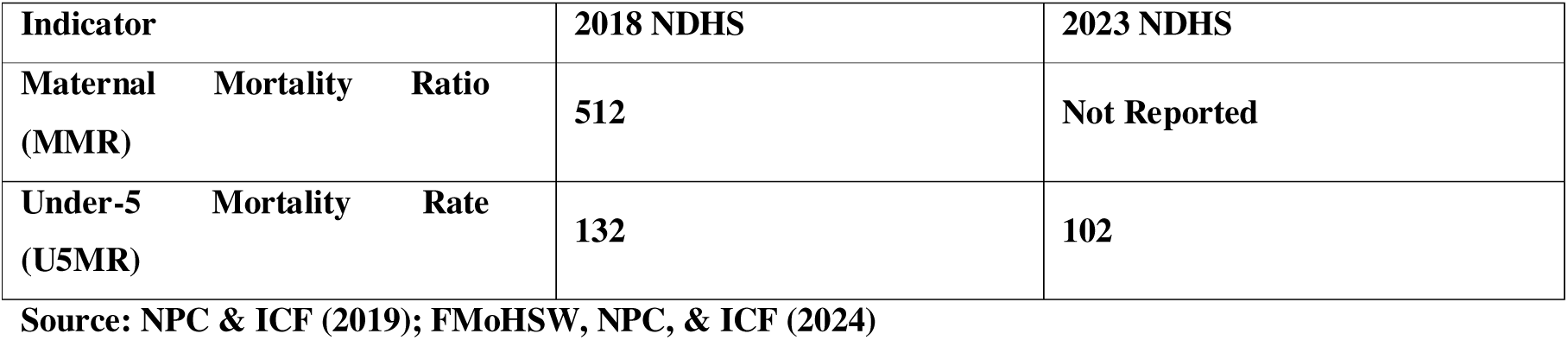
Maternal and Under-Five Mortality Indicators, 2018 vs 2023 NDHS.

**Figure 1:**
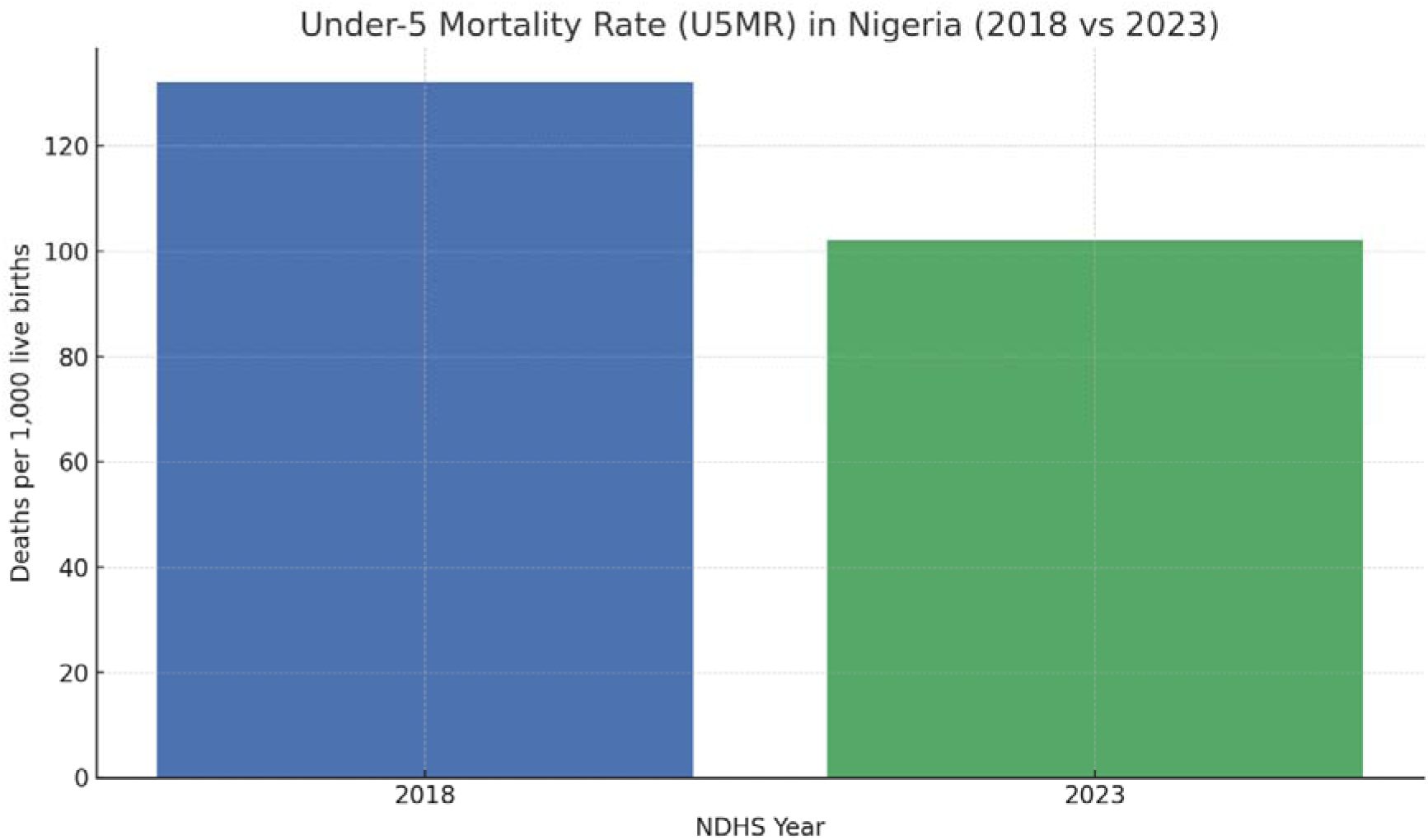
Under-5 Mortality Rate (U5MR) in Nigeria (2018 vs 2023) *Source: NPC & ICF (2019); FMoHSW, NPC, & ICF (2024)*

### 3.2 Immunization Coverage Trends (2018 vs 2023)

Between 2018 and 2023, Nigeria recorded moderate improvements in the national coverage of key childhood immunizations. According to the NDHS reports, the DPT3 vaccine coverage increased from 50% in 2018 to 57% in 2023, representing a 7-percentage-point improvement. The Measles vaccine coverage also rose from 54% to 59%, and Pentavalent vaccine coverage moved from 50% to 56% over the same period (NPC & ICF, 2019; FMoHSW, NPC, & ICF, 2024).

These gains suggest increased reach of routine immunization programmes, although national coverage still falls short of the ≥90% WHO target. Regional disparities remain a concern, with lower coverage in conflict-affected northern states compared to the south.

**Table 2:**
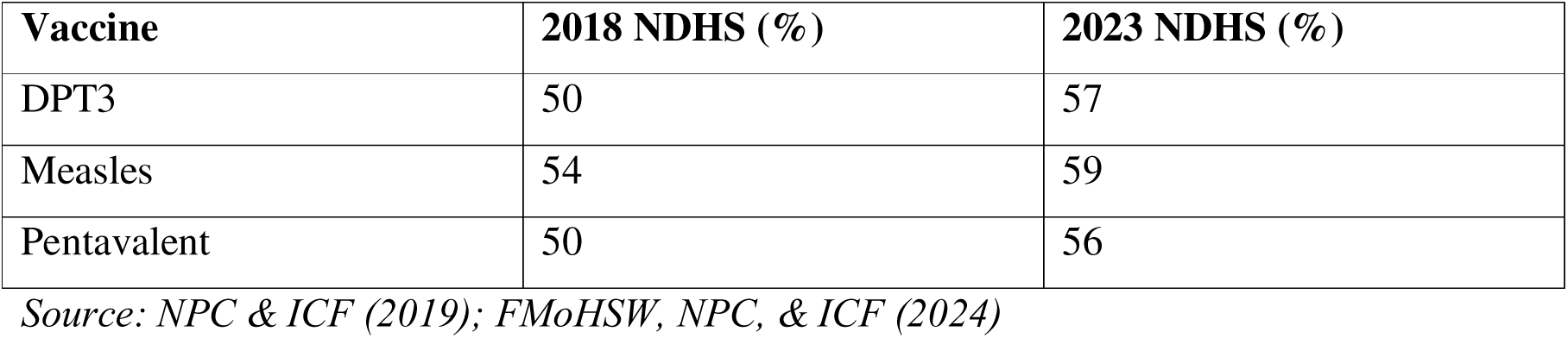
Immunization Coverage (%) by Vaccine Type, NDHS 2018 vs 2023.

**Figure 2:**
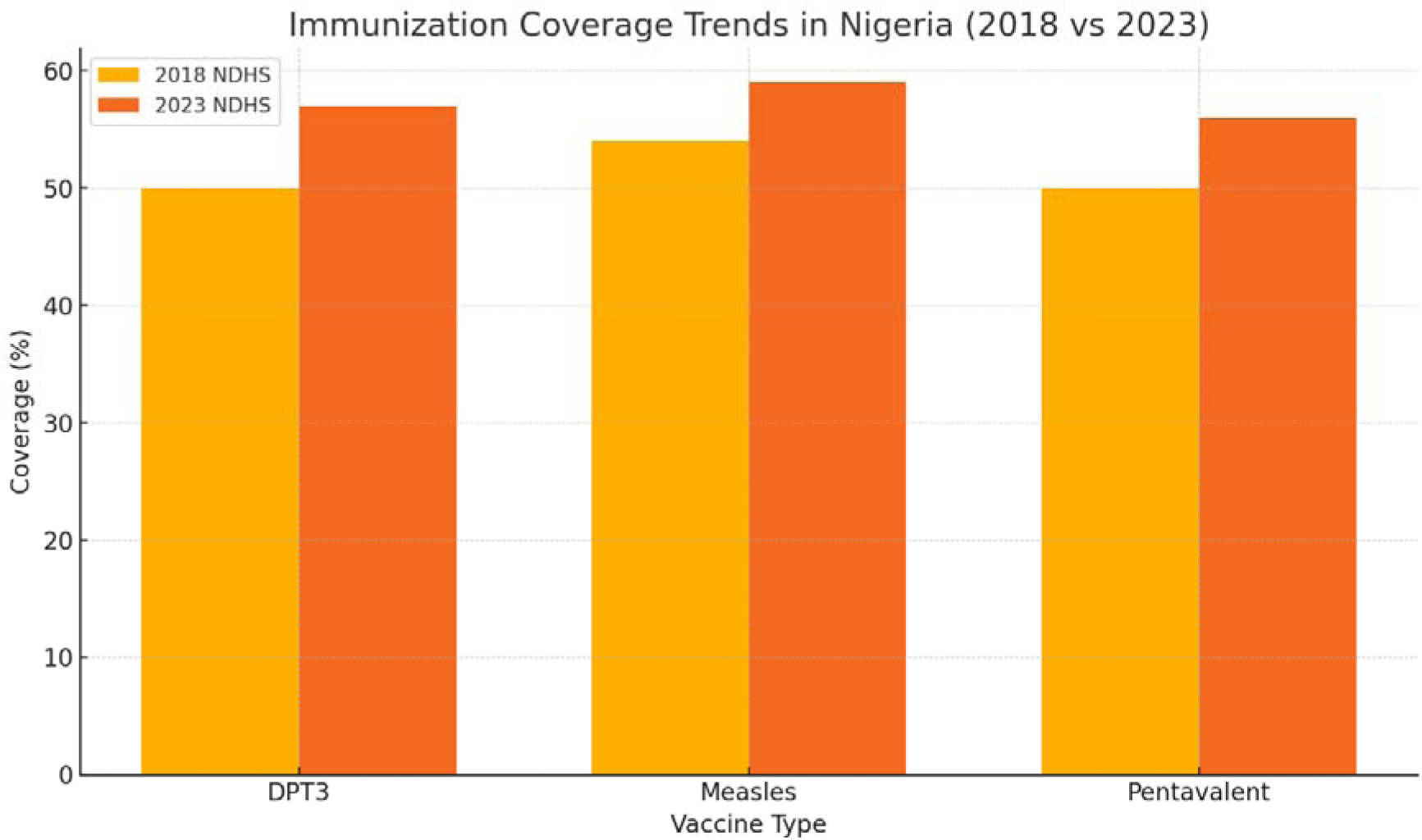
Immunization Coverage Trends in Nigeria (2018 vs 2023) *Source: NPC & ICF (2019); FMoHSW, NPC, & ICF (2024)*

### 3.3 Trends in Exclusive Breastfeeding and Malnutrition (2018 vs 2023)

The prevalence of exclusive breastfeeding among infants aged 0–5 months improved from 29% in 2018 to 34% in 2023, indicating better adherence to WHO/UNICEF recommendations on infant feeding practices. Progress was also observed across core malnutrition indicators: stunting reduced from 37% to 32%, wasting declined from 7% to 6%, and underweight children decreased from 22% to 19% over the five-year period (NPC & ICF, 2019; FMoHSW, NPC, & ICF, 2024).

These improvements suggest incremental progress in child nutrition, yet the figures remain above global targets and underscore the persistent challenge of undernutrition, especially in rural and food-insecure regions.

**Table 3:** Breastfeeding and Child Malnutrition Indicators, NDHS 2018 vs 2023.

| Indicator | 2018 NDHS (%) | 2023 NDHS (%) |
| --- | --- | --- |
| Exclusive Breastfeeding (0–5 months) | 29 | 34 |
| Stunting (Height-for-age) | 37 | 32 |
| Wasting (Weight-for-height) | 7 | 6 |
| Underweight (Weight-for-age) | 22 | 19 |

*Source: NPC & ICF (2019); FMoHSW, NPC, & ICF (2024)*

**Figure 3:**
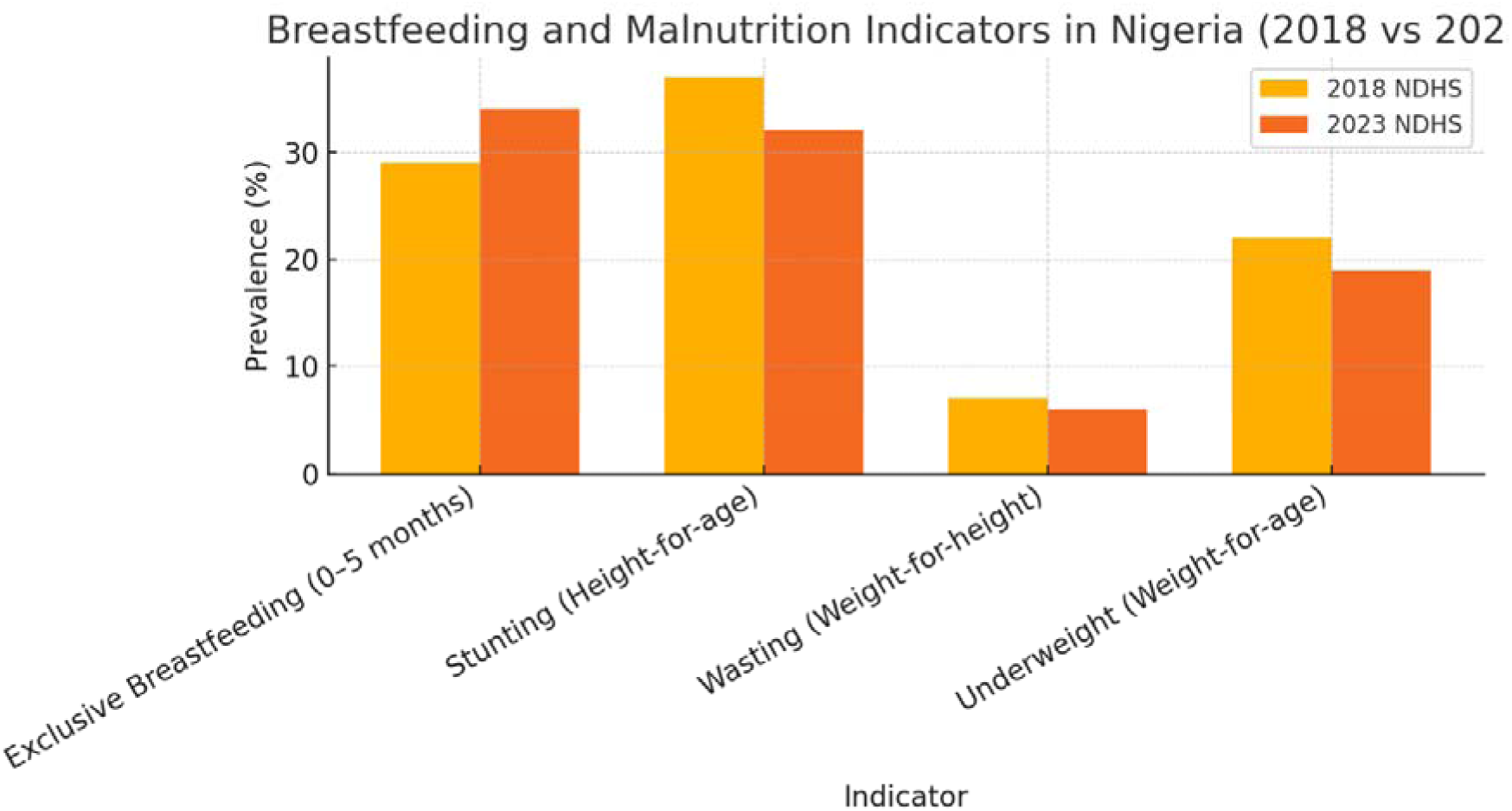
Breastfeeding and Malnutrition Indicators in Nigeria (2018 vs 2023) *Source: NPC & ICF (2019); FMoHSW, NPC, & ICF (2024)*

### 3.4 Antenatal Care, Skilled Birth Attendance, and Neonatal Survival (2018 vs 2023)

Between 2018 and 2023, maternal care indicators in Nigeria improved considerably. The proportion of women who received at least four antenatal care (ANC) visits increased from 57% to 68%, and skilled birth attendance (SBA) rose from 43% to 52%. Concurrently, the early neonatal mortality rate (ENMR) declined from 39 to 31 deaths per 1,000 live births, reflecting improvements in facility-based maternal and newborn care (NPC & ICF, 2019; FMoHSW, NPC, & ICF, 2024).

This trend supports global evidence linking increased ANC coverage and skilled delivery to better neonatal outcomes and lower early mortality risks.

**Table 4:**
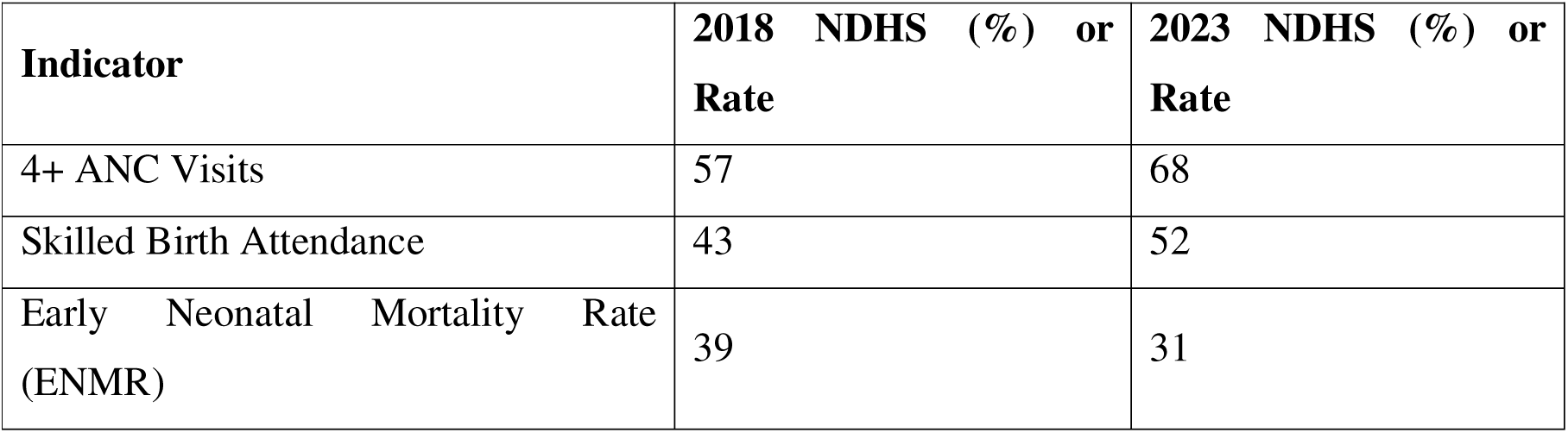
ANC, Skilled Attendance, and Early Neonatal Mortality (2018 vs 2023)

**Figure 4:**
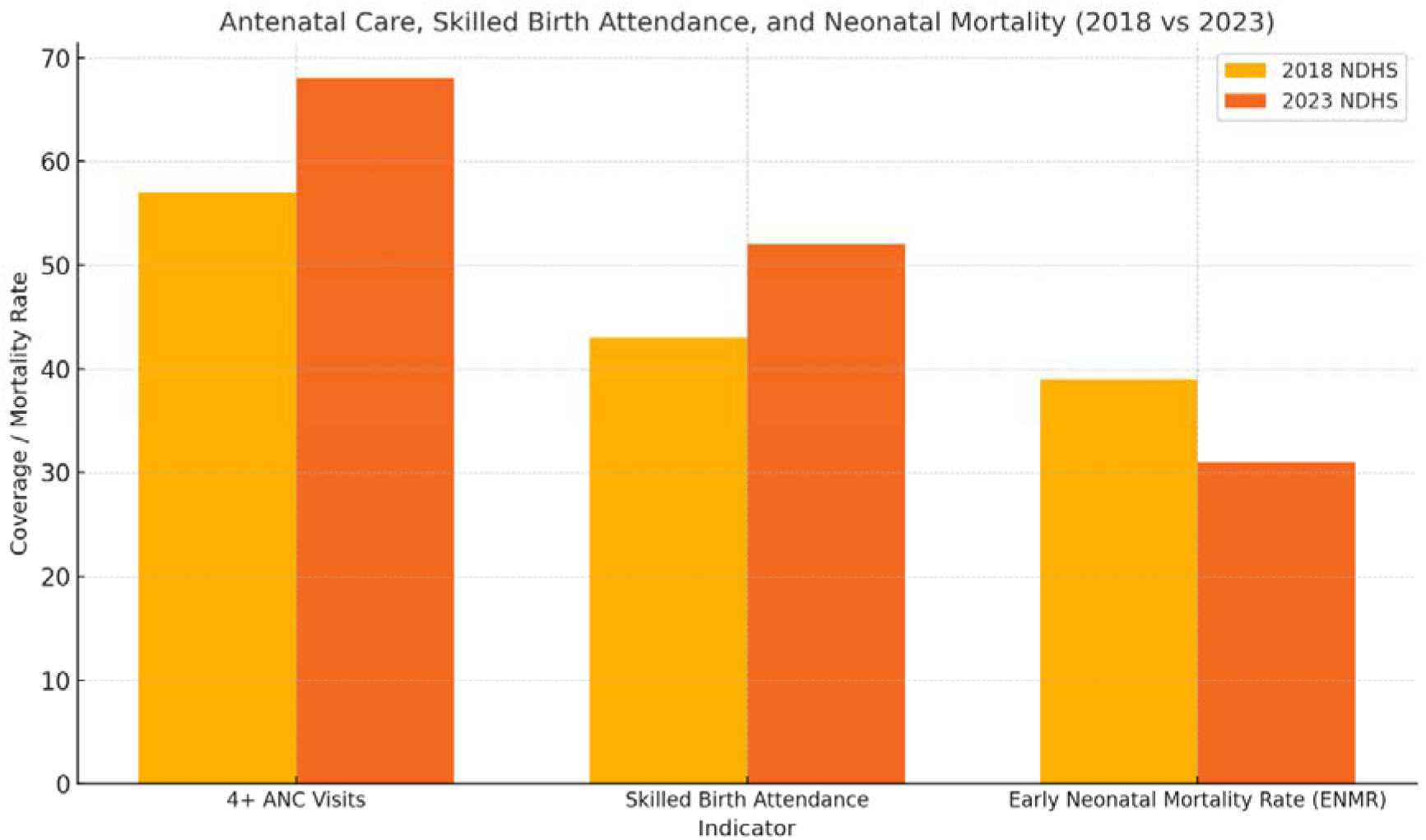
ANC, Skilled Birth Attendance, and Neonatal Mortality in Nigeria (2018 vs 2023)

### 3.5 Trends in Prevalence of Childhood Illnesses and Treatment-Seeking Behavior (2018 vs 2023)

Between 2018 and 2023, Nigeria recorded both declines in the prevalence of childhood illnesses and improvements in treatment-seeking behavior. The prevalence of fever in children under five dropped from 29% to 23%, while treatment sought for fever increased from 65% to 74%. Similarly, diarrhea prevalence decreased from 13% to 9%, and care-seeking for diarrhea rose from 43% to 53%. Symptoms of acute respiratory infection (ARI) declined marginally from 2% to 1%, with treatment-seeking increasing from 72% to 77% (NPC & ICF, 2019; FMoHSW, NPC, & ICF, 2024).

These changes reflect gains in community-based health awareness and improved access to pediatric healthcare, though regional gaps persist.

**Table 5:** Childhood Illnesses and Treatment-Seeking Trends, NDHS 2018 vs 2023.

| Indicator | 2018 NDHS (%) | 2023 NDHS (%) |
| --- | --- | --- |
| Fever in under-5s (Prevalence) | 29 | 23 |
| Treatment sought for fever | 65 | 74 |
| Diarrhea in under-5s (Prevalence) | 13 | 9 |
| Treatment sought for diarrhea | 43 | 53 |
| Symptoms of ARI (Prevalence) | 2 | 1 |
| Treatment sought for ARI | 72 | 77 |

*Source: NPC & ICF (2019); FMoHSW, NPC, & ICF (2024)*

**Figure 5:**
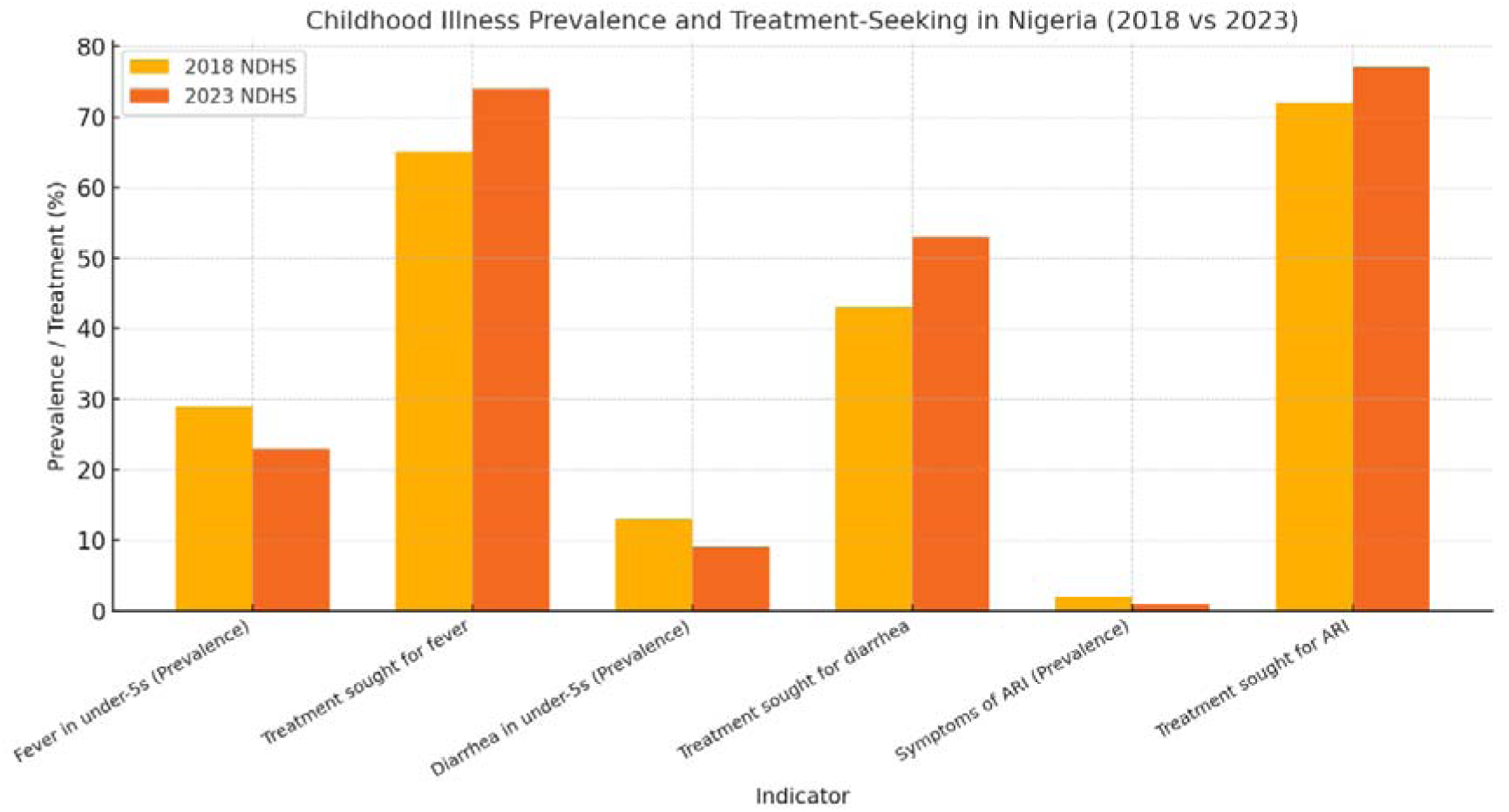
Childhood Illness Prevalence and Treatment-Seeking in Nigeria (2018 vs 2023)

### 3.6 Thematic Insights from NDHS Narrative Sections

The qualitative analysis using NVivo software revealed several prominent themes reflecting contextual barriers and enablers influencing maternal and child health in Nigeria between 2018 and 2023. Insights were derived from coding policy and narrative sections of the NDHS reports, focusing on access to services, health-seeking behavior, and system-level responsiveness. Four dominant themes emerged: geographic and infrastructural access, socioeconomic constraints, community-level trust and service engagement, and programmatic enablers.

#### 3.6.1 Geographic and Infrastructural Barriers

The most frequently coded references highlighted the impact of physical distance to health facilities and poor road infrastructure, especially in rural and conflict-affected zones. In multiple NDHS narrative sections, women reported difficulty accessing antenatal care (ANC) and skilled birth attendance due to the unavailability of nearby facilities or the cost of transportation. These structural constraints remain critical deterrents to timely service utilization, contributing to delays in seeking maternal care, as classified under the "first delay" model.

#### 3.6.2 Socioeconomic Constraints and Out-of-Pocket Costs

Another recurring theme was the effect of poverty and household-level financial insecurity on health-seeking behaviors. NDHS narratives revealed that out-of-pocket expenses for delivery supplies, medications, and transportation often discouraged women from attending ANC or choosing facility-based births. This finding was particularly pronounced in northern Nigeria and among households in the lowest wealth quintile. Despite national health insurance reforms, coverage remained limited, and cost remained a significant barrier for maternal and child health access.

#### 3.6.3 Trust, Community Engagement, and Service Uptake

Themes around community-level trust and engagement emerged as positive enablers. Respondents in several states noted increased confidence in health services where community health workers or facility-based staff were consistent, respectful, and locally rooted. The presence of Ward Health Committees and community volunteers was mentioned as having improved communication and mobilization, especially during immunization campaigns. Where trust was high, uptake of child immunization and early ANC initiation also increased.

#### 3.6.4 Programmatic Enablers and Gaps

While coding policy-related NDHS content, instances were identified where government programs such as the Basic Health Care Provision Fund (BHCPF) and free maternal and child health schemes were described as positively impacting service utilization. However, these enablers were inconsistently implemented. Some respondents noted frequent stock outs, poorly staffed facilities, or referral failures, which negated the potential benefits of such programs. The gap between policy promise and frontline experience was a cross-cutting theme in the narrative data.

## 4.0 DISCUSSION

### 4.1 Maternal Mortality Ratio (MMR) and Under-5 Mortality Rate (U5MR)

The comparative analysis of the Nigeria Demographic and Health Surveys (NDHS) from 2018 and 2023–24 illustrates a measurable improvement in child survival, particularly with regard to the under-five mortality rate (U5MR). According to the NDHS 2018, U5MR stood at 132 deaths per 1,000 live births. By the time of the 2023–24 survey, this figure had declined to 102 deaths per 1,000 live births. This represents an estimated 22.7% reduction over a five-year period a substantial gain within the Nigerian context where multiple socio-economic and health system challenges persist. The reduction, while encouraging, still falls short of the Sustainable Development Goal (SDG) 3.2, which aims to reduce U5MR to fewer than 25 deaths per 1,000 live births by 2030. Nevertheless, the downward trend provides evidence of gradual progress toward improving child survival outcomes in Nigeria. Multiple factors are likely responsible for this improvement. First, the expansion of routine immunization coverage across several regions of the country has likely contributed to lower child mortality. As detailed in related findings, increased coverage of vaccines such as DPT3, measles, and the pentavalent combination vaccine reduces susceptibility to common and often fatal childhood illnesses. Immunization has a proven effect not only in direct disease prevention but also in fostering broader health-seeking behavior among caregivers, reinforcing contact between families and healthcare providers.

In parallel, advancements in maternal health services have likely had indirect yet meaningful effects on neonatal and child survival. Improvements in antenatal care (ANC) utilization, skilled birth attendance (SBA), and postnatal care may have resulted in fewer birth complications, timely detection of pregnancy-related risks, and better post-delivery support for both mothers and newborns. Such services are crucial in reducing the incidence of neonatal conditions that can contribute to early childhood mortality, including infections, birth asphyxia, and low birth weight. Additionally, growing awareness of child nutrition, exclusive breastfeeding, and early infant care has become more widespread through targeted outreach programs and community-based interventions. Nigeria’s participation in initiatives like the Integrated Management of Childhood Illness (IMCI) strategy may also have had a positive impact. This approach emphasizes not only curative care but also disease prevention, nutritional support, and education for caregivers, with a focus on improving the diagnosis and treatment of pneumonia, diarrhea, malaria, and malnutrition. The scaled-up distribution of insecticide-treated nets (ITNs), deworming campaigns, and the promotion of oral rehydration therapy (ORT) have helped strengthen community-level resilience to common health threats that disproportionately affect children.

Despite these improvements, the pace of change remains slow when juxtaposed with global targets. A U5MR of 102 still places Nigeria among the countries with the highest child mortality rates worldwide. This reality points to underlying structural challenges in Nigeria’s health system that continue to limit the scale and impact of interventions. Notably, the national U5MR figure conceals wide disparities across geographic regions, socioeconomic strata, and urban-rural divides. For instance, children in northern Nigeria continue to face a much higher risk of dying before age five compared to those in southern zones. These differences are influenced by factors such as maternal education, household income, cultural norms, and accessibility to healthcare services.

The risk gradient becomes even steeper when intersecting vulnerabilities are considered. Children from rural, low-income households with mothers who have limited formal education are far less likely to be vaccinated, receive prompt medical care during illness, or be delivered in a facility with skilled attendants. Additionally, infrastructure challenges such as poor roads, limited electricity, and inadequate water supply in remote areas make it difficult for families to reach care and for health providers to offer consistent services. As a result, while national statistics may reflect general progress, large segments of the population continue to experience disproportionately high mortality risks. While data on U5MR has been updated in the 2023–24 Key Indicators Report, a major limitation in evaluating progress on maternal health is the absence of an updated maternal mortality ratio (MMR) figure. The 2018 NDHS reported an MMR of 512 deaths per 100,000 live births, a figure that signaled deep systemic gaps in emergency obstetric care, particularly in terms of facility preparedness, skilled health personnel, and timely referral systems. The lack of a comparable 2023–24 estimate prevents a full understanding of whether maternal health outcomes have followed a trajectory similar to child health. There are several plausible reasons for the omission of updated MMR data in the preliminary 2023–24 release. Estimating maternal mortality is methodologically complex, often requiring large sample sizes and sophisticated modeling techniques to account for the relative rarity of maternal deaths and potential misclassification. These estimates are usually presented in final DHS reports or sourced from international modeling groups such as the Maternal Mortality Estimation Inter-agency Group (MMEIG). Until the final NDHS report is published or supplementary sources provide an updated national MMR, assessments of Nigeria’s progress toward SDG 3.1 which targets a reduction to fewer than 70 maternal deaths per 100,000 live births will remain speculative.

Nevertheless, child mortality trends often reflect the broader performance of the health system. A decline in U5MR implies that at least some components of the system particularly at the primary care level are improving. These may include better outreach services, increased use of community health workers, and more consistent delivery of preventive and curative care. At the same time, it is crucial to recognize that gains in child survival do not automatically translate to parallel improvements in maternal health. Reducing maternal mortality requires specific investments in emergency obstetric care, including access to skilled personnel, safe surgical capabilities for cesarean sections, access to blood transfusions, and timely management of conditions such as eclampsia and postpartum hemorrhage.

The linkage between maternal and child health outcomes, while conceptually strong, must be operationalized through integrated programming and service delivery. For example, ANC visits should be used not only to monitor fetal growth and maternal blood pressure but also to educate mothers about newborn care, immunization schedules, nutrition, and family planning. Similarly, postnatal services should cover both the health of the mother and the newborn to ensure comprehensive early-life care. The improvements in U5MR observed in Nigeria between 2018 and 2023 reflect important public health achievements but also reveal areas where progress remains fragile and uneven. Structural inequalities, data gaps, and the continued absence of updated maternal mortality data limit the extent to which these gains can be understood and built upon.

### 4.2 Trends in Child Immunization Coverage

The trends in child immunization coverage in Nigeria, as captured by the 2018 and 2023 Nigeria Demographic and Health Surveys (NDHS), demonstrate a modest yet meaningful progression in the nation’s efforts to protect children from vaccine-preventable diseases. Over the five-year period, measurable gains were recorded across key immunization indicators. The coverage for the third dose of the DPT vaccine (DPT3) improved from 50% in 2018 to 57% in 2023. Similarly, measles vaccination coverage rose from 54% to 59%, while pentavalent vaccine coverage increased from 50% to 56%. These developments, while modest in scale, reflect a slowly strengthening immunization system responding to national and global pressure to reduce child mortality and morbidity.

Immunization is one of the most cost-effective public health interventions and remains a cornerstone of child survival strategies. The upward trends in vaccine coverage suggest an ongoing expansion of service delivery, likely supported by more extensive outreach programming and targeted interventions in previously underserved populations. The increased figures, though not dramatic, are significant when viewed within the context of Nigeria’s complex health landscape marked by vast geographic, socioeconomic, and infrastructural disparities. One of the plausible drivers of the observed improvements is the strengthening of immunization campaigns at the subnational level. State governments, often in partnership with international organizations and non-governmental agencies, have increasingly adopted strategies such as routine immunization (RI) intensification days, house-to-house mobilization, and the use of trained community health volunteers. These activities have enhanced awareness, improved logistics, and encouraged more timely presentation of children for vaccinations.

Mobile vaccination campaigns have also been particularly influential, especially in hard-to-reach areas. The use of temporary immunization posts in schools, marketplaces, and religious centers has contributed to reaching populations in remote or conflict-affected regions. Similarly, innovations such as the deployment of solar-powered vaccine refrigerators and GPS-enabled logistics tracking have helped to maintain the cold chain in environments previously thought to be unserviceable. These innovations are crucial in a country where vast distances, unreliable electricity, and seasonal weather patterns present major logistical challenges.

Nonetheless, it is important to acknowledge that despite these efforts, the national immunization coverage still falls significantly below the World Health Organization’s (WHO) recommended threshold of 90% required to achieve herd immunity. This shortfall exposes many Nigerian children to continued risk of preventable illnesses such as diphtheria, pertussis, tetanus, and measles. Outbreaks of these diseases are not uncommon, even in the presence of ongoing immunization programs, due largely to suboptimal coverage in key districts and local government areas. A major barrier to achieving higher immunization coverage is the persistent inequity between rural and urban areas. Children living in urban centers generally have higher access to health facilities and more consistent exposure to health messaging through media and mobile communication. In contrast, rural children often live in communities with inadequate infrastructure, limited healthcare personnel, and long distances to service points. Insecurity in parts of the northeast and northwest further compounds these challenges, as health workers are sometimes unable to operate safely or are forced to reduce the frequency and scope of outreach.

Misinformation and vaccine hesitancy also continue to hamper efforts, particularly in certain northern regions where past resistance to polio vaccination campaigns still lingers. Distrust of government services, influenced by political instability and historical neglect, has translated into skepticism about immunization safety and efficacy. This is worsened by the proliferation of disinformation through unregulated social media platforms. Some communities have also been influenced by religious or cultural misconceptions that discourage engagement with health services, including immunization. Even in communities where awareness exists, structural weaknesses within the health system often impede effective service delivery. Long waiting times at health facilities, stock outs of vaccines, and unavailability of trained personnel all contribute to missed opportunities. In some cases, caregivers are turned away due to the lack of necessary supplies or asked to return on multiple occasions, which discourages completion of vaccination schedules.

There is also a significant gap in the quality of data reporting and monitoring. Incomplete or inaccurate recording of vaccinations has led to underestimation of true coverage levels in some regions and overestimation in others. This undermines planning and resource allocation. Without accurate real-time data, it becomes difficult to track children who are under-vaccinated or completely missed. Paper-based systems, though still widely used, are increasingly being recognized as insufficient for managing complex immunization data, especially in high-volume or mobile populations.

To address these challenges, several capacity-building and digital initiatives have been introduced in select states. The integration of electronic immunization registries, mobile health (mHealth) tracking tools, and dashboard monitoring has begun to yield dividends in terms of data quality and timely response. Digital systems enable health workers to flag missed vaccinations, schedule follow-up reminders, and improve accountability for vaccine usage. However, the scale-up of such technologies remains uneven across the country and is heavily dependent on donor funding and local political will.

Another often-overlooked challenge is the burden placed on caregivers. In many households, the responsibility of taking a child for vaccination falls entirely on women, who may be constrained by time, finances, or the absence of decision-making autonomy within the household. A lack of support from male partners or extended family members can reduce the likelihood that a child completes the full immunization schedule. Community mobilization strategies that engage men and elders as well as religious and traditional leaders have been identified as successful models for improving uptake.

Supply chain stability remains a critical determinant of vaccine coverage. Nigeria’s immunization supply chain is vast and complex, involving multiple storage levels, transport logistics, and coordination between federal and state agencies. Any breakdown in this chain whether due to fuel shortages, budget delays, or procurement inefficiencies can lead to missed vaccinations. Strengthening the supply chain system with modern tools, efficient forecasting, and cold chain expansion will be essential for ensuring continuous availability of vaccines at all levels of the health system.

There is also a growing need to transition from vertical, campaign-style delivery models to integrated, people-centered approaches. Integrating immunization services with maternal and child health programs, nutrition screening, and growth monitoring has the potential to improve both uptake and efficiency. Families are more likely to access services when multiple needs can be met in a single visit, reducing transportation costs and time lost from daily responsibilities.

Despite the ongoing challenges, the steady increase in vaccine coverage between 2018 and 2023 should be acknowledged as a positive trend. The gains achieved demonstrate that when immunization is prioritized through coordinated multi-level engagement from government to community level improvements are possible. However, sustaining and accelerating this progress will require more than just technical adjustments. It will demand a deliberate focus on equity, data-driven decision-making, reliable financing, and an unwavering commitment to health system strengthening.

### 4.3 Exclusive Breastfeeding and Malnutrition Indicators

The results from the 2018 and 2023 NDHS reports reveal a mixed but encouraging picture in terms of infant feeding practices and child nutrition outcomes. Exclusive breastfeeding (EBF) for infants aged 0–5 months increased from 29% to 34%, a positive sign of growing adherence to WHO and UNICEF recommendations. Over the same period, the prevalence of child stunting declined from 37% to 32%, wasting decreased slightly from 7% to 6%, and underweight rates dropped from 22% to 19%.

The upward movement in exclusive breastfeeding rates may be attributed to expanded health education efforts, community-level counseling, and baby-friendly hospital initiatives that have been adopted in parts of Nigeria. These improvements reflect gradual changes in maternal knowledge and behavior regarding the importance of breastfeeding in the first six months of life. Still, the coverage remains well below the global target of 50%, suggesting significant room for scale-up.

The decline in malnutrition indicators, particularly stunting and underweight prevalence, points to the impact of integrated nutrition programs and improved food access in some regions. However, the persistent rates indicate that malnutrition remains a public health concern. The regional disparities in nutritional outcomes remain stark, with higher rates of stunting and wasting in northern states, where food insecurity, limited healthcare access, and poverty are more acute.

Factors contributing to persistent malnutrition include suboptimal complementary feeding, recurrent infections, low maternal education, and poor sanitation. Although progress has been made, many children still do not receive the minimum acceptable diet required for optimal growth and development. Household poverty and rising food prices have also posed new challenges to dietary quality and diversity.

Moreover, structural and systemic factors such as the limited reach of nutrition services, low coverage of growth monitoring programs, and a shortage of trained nutrition professionals have restricted the effectiveness of national nutrition efforts. The burden of malnutrition also places long-term pressures on the health and education systems, given its association with delayed cognitive development and increased susceptibility to illness. Addressing these gaps requires a multisectorial approach that includes agriculture, water and sanitation, social protection, and health. Community-based management of acute malnutrition (CMAM), cash transfer programs, and targeted food supplementation schemes can serve as catalytic tools for progress if properly coordinated and sustained. Nigeria has achieved modest gains in both breastfeeding practices and malnutrition reduction between 2018 and 2023. However, the prevalence of under nutrition remains high and progress is uneven. To meet global nutrition targets, efforts must shift from sporadic programmatic interventions to more sustainable, equity-focused national strategies.

### 4.4 Antenatal Care, Skilled Birth Attendance, and Neonatal Survival

The comparative data from the 2018 and 2023 NDHS reports demonstrate a steady improvement in Nigeria’s maternal healthcare indicators, particularly in antenatal care coverage and skilled birth attendance. The proportion of women who received four or more antenatal care (ANC) visits increased from 57% in 2018 to 68% in 2023, while skilled birth attendance (SBA) rose from 43% to 52% over the same period. Concurrently, the early neonatal mortality rate (ENMR) defined as the number of deaths during the first seven days of life per 1,000 live births declined from 39 to 31.

This progression, though incremental, is significant for a country like Nigeria where maternal and neonatal mortality remain among the highest in the world. The trends observed support the widely documented association between increased maternal health service utilization and improved neonatal outcomes. Antenatal care serves as a critical entry point for pregnant women into the health system, allowing for early detection of complications, management of pre-existing conditions, and administration of preventive interventions such as tetanus immunization, iron and folate supplementation, and intermittent preventive treatment for malaria in pregnancy. The increase in ANC utilization could be attributed to a combination of policy and programmatic interventions undertaken in recent years. These include community health outreach campaigns, conditional cash transfer initiatives, and awareness programs advocating for early and consistent ANC attendance. Additionally, the increased coverage of health facilities offering ANC services, including those integrated into primary healthcare centers, has likely contributed to this improvement. More pregnant women are also receiving care from trained health personnel, including midwives and nurses, rather than relying solely on traditional birth attendants.

However, while the rise in the percentage of women attending four or more ANC visits is encouraging, it is important to note that the quality and timing of these visits are equally important. Many women still initiate ANC late in their pregnancies, often in the second or third trimester, which reduces the opportunity for early risk identification and intervention. Moreover, not all ANC visits are comprehensive some may lack essential diagnostic procedures or counseling components due to resource constraints or staffing issues. Hence, increasing coverage must go hand in hand with ensuring that the content and delivery of ANC meet minimum quality standards.

The growth in skilled birth attendance is another promising development. The shift from 43% to 52% between 2018 and 2023 suggests that more women are now giving birth in health facilities or are attended to by trained healthcare providers during delivery. Skilled attendance at birth is one of the most effective strategies for reducing both maternal and neonatal mortality, as it ensures the timely management of complications such as postpartum hemorrhage, birth asphyxia, and obstructed labor.

Nevertheless, this progress is tempered by persistent gaps in access and equity. Rural women, particularly in the northern regions of Nigeria, continue to face numerous barriers to facility-based deliveries, including distance, cost, cultural norms, and lack of transport. Insecurity in certain parts of the country also deters women from seeking care in formal health settings. Urban-rural and regional disparities remain stark, with skilled birth attendance often exceeding 70% in urban areas while falling below 30% in some rural communities.

Despite these challenges, the observed decline in the early neonatal mortality rate from 39 to 31 deaths per 1,000 live births offers strong evidence that improvements in maternal health service delivery are beginning to translate into better outcomes for newborns. Early neonatal deaths are primarily caused by complications during childbirth, including prematurity, intrapartum-related events (commonly known as birth asphyxia), and neonatal infections. Reducing these deaths requires a continuum of care approach beginning with effective ANC, continuing through skilled delivery, and extending into the early postnatal period.

The reductions in ENMR could, therefore, be reflective of better intrapartum management and increased uptake of neonatal care services. In facilities where skilled attendants are present, timely resuscitation of newborns, proper cord care, immediate and exclusive breastfeeding, and infection prevention practices are more likely to be implemented. The presence of trained healthcare workers also facilitates quick referral for babies showing signs of distress, which is crucial in the critical first days of life.

Another factor potentially influencing neonatal survival is the rising integration of maternal and newborn health programs. The increased use of partographs to monitor labor, improved infection control protocols, and expanded training for midwives and community health extension workers have likely contributed to improved outcomes. National strategies such as the Nigeria Every Newborn Action Plan (NENAP) have also laid down critical policy frameworks to support reductions in neonatal mortality.

That said, sustaining and expanding these gains will require deliberate investment and system-wide reforms. There is still a shortage of skilled health professionals, particularly in rural areas, and many facilities lack basic supplies such as clean delivery kits, magnesium sulfate, and neonatal resuscitation equipment. The gaps in infrastructure, including inadequate water and sanitation in health centers, also compromise maternal and newborn outcomes.

Moreover, poor health information systems and underreporting continue to hinder effective tracking of maternal and neonatal outcomes. A significant portion of births and neonatal deaths, particularly those occurring at home, are not recorded in the formal system. Strengthening civil registration and vital statistics (CRVS) systems is therefore essential for better monitoring and planning. From a policy perspective, the findings highlight the importance of sustained focus on maternal and newborn health within Nigeria’s broader universal health coverage agenda. Subsidizing maternal health services, improving transportation and referral systems, and integrating birth preparedness counseling during ANC could further enhance service uptake. Additionally, male involvement in maternal health and continued community engagement will be key to shifting social norms that hinder facility-based deliveries.

### 4.5 Childhood Infectious Diseases and Treatment-Seeking Behavior

Childhood infectious diseases particularly malaria, diarrhea, and acute respiratory infections (ARI) remain leading causes of morbidity and mortality among children under five in Nigeria. The NDHS 2018 and 2023–24 comparison provides critical insights into the evolving epidemiological trends of these diseases and, more importantly, into the health-seeking behavior of caregivers in response to childhood illness.

The data show a significant reduction in the prevalence of fever (often used as a proxy indicator for malaria) from 29% in 2018 to 23% in 2023 among under-five children. Similarly, diarrhea prevalence declined from 13% to 9%, and symptoms of ARI fell slightly from 2% to 1%. These changes suggest a positive trend in the burden of childhood infectious diseases, likely influenced by public health interventions such as improved vaccination coverage, increased use of insecticide-treated nets (ITNs), improved sanitation practices, and rising awareness among caregivers.

Malaria control, in particular, has benefitted from robust global and national efforts. Insecticide-treated net distribution campaigns, community-based education on mosquito prevention, and intermittent preventive treatment during pregnancy (IPTp) have contributed to reductions in malaria transmission among children. Furthermore, the integration of malaria diagnosis and treatment at the primary healthcare level has made access to care more efficient. The reduction in fever prevalence could reflect both a real decline in disease burden and improved preventive behavior among households.

The decrease in diarrhea prevalence is also encouraging. This may be attributed to better water, sanitation, and hygiene (WASH) practices. The increasing availability of clean drinking water, hand washing stations, and public health messaging on safe food handling likely contributed to this outcome. Similarly, the decrease in ARI symptoms though modest may reflect gains in immunization coverage, reduction in household air pollution (from improved cooking methods), and better nutritional outcomes for children, which contribute to stronger immune defenses.

While reductions in disease prevalence are important, the corresponding increase in treatment-seeking behavior is equally notable. Between 2018 and 2023, the proportion of caregivers who sought treatment for children with fever rose from 65% to 74%. For diarrhea, care-seeking increased from 43% to 53%, and for ARI, it improved from 72% to 77%. These improvements indicate an encouraging shift toward timely and appropriate health service utilization during episodes of childhood illness.

This trend may be due in part to growing public awareness and the expansion of primary health centers across the country. Health campaigns promoting early treatment, the introduction of free or subsidized child health services in certain states, and increased female education known to be positively correlated with healthcare-seeking may have played pivotal roles. Moreover, mobile outreach clinics and the role of community health workers (CHWs) in identifying and referring sick children have strengthened Nigeria’s frontline healthcare system.

Despite these improvements, significant barriers to treatment still persist, particularly in rural and conflict-affected areas. In many such settings, geographic distance to healthcare facilities, lack of transportation, high out-of-pocket costs, and deeply rooted cultural beliefs continue to hinder optimal care-seeking. In some communities, caregivers still prefer traditional remedies or delay seeking care due to financial constraints or lack of decision-making power among women. These systemic and sociocultural constraints remain a formidable challenge to achieving universal coverage of essential child health services.

Additionally, although more caregivers are seeking treatment, questions remain about the **quality** of care received. There is evidence that many children who are brought to health facilities do not receive a full clinical assessment or the appropriate medications. Stock outs of essential drugs (e.g., antimalarials, oral rehydration salts, antibiotics), limited diagnostic capacity, and the overreliance on unqualified drug vendors and patent medicine shops undermine the potential benefits of increased treatment-seeking.

Another concern is the use of health services from informal or unregulated providers. While many caregivers may seek help for their children’s illness, they may do so at outlets that do not adhere to standard clinical guidelines. This results in inappropriate treatment, misuse of antibiotics, or delayed referral of severe cases. Ensuring that care-seeking translates into effective and safe treatment will require better training, regulation, and supervision of all child health service providers, including those operating at the community level.

Moreover, the differences in care-seeking behavior by region, maternal education, and household wealth quintiles are still pronounced. Children from wealthier households and those whose mothers have at least secondary education are far more likely to be taken to a formal health provider. These disparities highlight the need for equity-focused strategies in child health programming. Targeted subsidies, demand-side financing (e.g., conditional cash transfers), and community-based insurance models could help bridge this gap.

On the supply side, ensuring the functionality of primary health centers is critical. Many facilities in rural Nigeria suffer from staffing shortages, erratic power supply, limited diagnostic tools, and poor water and sanitation. Without addressing these bottlenecks, the gains in treatment-seeking may not be sustainable. Strengthening the operational capacity of frontline health facilities is therefore central to improving both access and outcomes.

From a policy and planning perspective, these findings support the continued prioritization of child health within Nigeria’s broader health system reform agenda. Integration of child health services into routine outreach campaigns, linking child survival efforts with nutrition and maternal care programs, and enhancing community-level health governance structures will be important for accelerating progress. The use of mobile health (mHealth) tools to remind caregivers about follow-up visits or deliver health education messages also offers new opportunities to improve service uptake.

Finally, it is essential to emphasize that while prevalence rates have declined and treatment-seeking has increased, the progress is uneven and, in some cases, fragile. Insecurity, economic shocks, and climate-related disasters could disrupt service delivery and reverse the gains made. For instance, flooding in northern Nigeria has displaced thousands of households and disrupted routine healthcare services in affected communities, increasing the risk of outbreaks and reducing access to care. To safeguard progress, child health interventions must be embedded within resilient health systems capable of withstanding shocks. Strengthening surveillance systems, improving supply chain resilience, and investing in health workforce development are all crucial to protecting the health of Nigeria’s youngest and most vulnerable citizens.

### 4.6 Thematic Interpretation of Maternal and Child Health Outcomes

The NVivo-assisted qualitative analysis of policy and narrative sections within the NDHS reports offers rich contextual insights that complement the quantitative data and deepen the understanding of Nigeria’s progress toward maternal and child health improvement between 2018 and 2023. While numeric indicators such as under-five mortality, immunization coverage, exclusive breastfeeding, and skilled birth attendance show modest improvement, the qualitative findings illuminate the lived experiences, structural challenges, and enabling factors that shape these trends on the ground. These insights directly reinforce and interpret the five core objectives of the study.

The reported reduction in under-five mortality from 132 to 102 deaths per 1,000 live births appears to reflect gradual improvements in child survival interventions. However, NDHS narrative accounts consistently emphasized disparities in access particularly in rural and conflict-affected communities where long distances to health facilities, poor road infrastructure, and limited emergency transportation delay or prevent timely care-seeking. These geographic and infrastructural challenges contribute to adverse outcomes not only for child health but also for maternal survival, highlighting how access remains unevenly distributed across Nigeria’s regions. The inability to re-evaluate maternal mortality trends in the 2023 NDHS further underscores systemic limitations in data completeness and real-time surveillance, thereby obstructing accountability on SDG 3.1.

Immunization coverage for key childhood vaccines such as DPT3, measles, and pentavalent improved marginally during the review period. Yet, qualitative responses indicate that vaccine hesitancy, caregiver mistrust, and service inconsistencies still constrain full immunization. In areas where caregivers reported positive interactions with local health workers especially those rooted in the community uptake was notably higher. Conversely, communities where staff were absent, poorly trained, or disrespectful experienced persistent resistance to vaccination efforts. These findings affirm the critical role of trust, community engagement, and interpersonal service quality in achieving immunization targets.

Exclusive breastfeeding rates increased from 29% to 34%, an achievement attributed in part to increased facility-based counseling. However, NDHS narratives also revealed that misinformation, low maternal literacy, peer pressure, and early return to work disrupted breastfeeding continuity. Reports from mothers indicated that malnutrition in children was often linked to maternal dietary insufficiency and economic hardship. Stunting and underweight prevalence were particularly associated with households in the lowest income quintiles and among mothers with limited education. These findings suggest that while health facility interventions help promote positive feeding practices, sustainable nutrition outcomes require integrated social and economic support mechanisms.

Improved access to antenatal care and skilled birth attendance reflected by increased ANC (4+ visits) and a rise in institutional deliveries was shaped by both programmatic support and community dynamics. However, NDHS narratives also noted delayed ANC initiation due to indirect costs such as transportation, medications, and informal facility charges. Women frequently cited overcrowding, long wait times, and poor staff attitudes as deterrents to returning for follow-up visits or opting for facility-based delivery. In contrast, areas where Ward Health Committees were active and community health volunteers were engaged reported higher levels of ANC completion and institutional births. These findings reinforce that maternal service uptake is influenced as much by perceptions of care quality and local engagement as by service availability.

Although treatment-seeking for childhood illnesses such as fever, diarrhea, and acute respiratory infections improved, the narrative sections revealed variations in the type and timeliness of care accessed. Caregivers in rural or low-resource settings often delayed seeking formal care due to cost or distance and turned first to traditional remedies. However, the presence of accessible, well-known community health workers boosted caregiver confidence and encouraged earlier treatment-seeking. These insights underscore the importance of frontline worker presence and culturally aligned care approaches in improving child health outcomes.

Across all objectives, one of the most persistent cross-cutting themes was the importance of trust in the health system. Communities that trusted their health providers were more likely to engage with services, complete immunization schedules, and deliver at health facilities. However, where trust had been eroded through stock outs, neglect, or disrespect communities either disengaged from the formal system or sought care elsewhere. These findings highlight that technical interventions must be paired with consistent and respectful provider behavior to succeed.

Another major insight from the narrative data was the inconsistency in program implementation. National programs such as the Basic Health Care Provision Fund (BHCPF) and free maternal and child health schemes were mentioned across regions but with wide variability in delivery. In some areas, services were described as well-organized and impactful. In others, frequent stock outs, understaffing, and lack of follow-up limited their effectiveness. This variation speaks to deeper governance and accountability gaps that undermine national policy execution.

Lastly, the analysis confirmed the pressing issue of weak data systems. While the NDHS provides a comprehensive overview of national health trends, the delay in releasing full survey reports and the absence of disaggregated data by state or local government area limit the utility of such data for real-time planning and intervention. Without timely, granular information, local health authorities struggle to respond adaptively to region-specific needs or monitor the equity of service distribution.

In sum, the qualitative evidence from the NDHS narrative sections strongly supports the study’s quantitative findings, while offering a richer understanding of the socio-cultural, systemic, and community-based factors influencing maternal and child health outcomes in Nigeria. These insights emphasize the importance of contextualized, people-centered approaches, not just to expand service coverage but to ensure that services are trusted, accessible, and equitably delivered across all communities.

## 5.0 CONCLUSIONS

This research project was designed to provide a comprehensive evaluation of maternal and child health trends in Nigeria between 2018 and 2023, with a particular focus on assessing the country’s progress toward Sustainable Development Goals (SDGs) 3.1 and 3.2. By systematically comparing nationally representative data from the Nigeria Demographic and Health Surveys (NDHS) 2018 and 2023–24, and augmenting this with contextual qualitative analysis using NVivo software, the study offers both empirical and thematic perspectives on Nigeria’s maternal and child health outcomes.

The study was anchored on five key objectives: to compare maternal and under-five mortality rates; assess immunization coverage; examine exclusive breastfeeding and malnutrition indicators; evaluate trends in antenatal care (ANC) and skilled birth attendance (SBA); and explore treatment-seeking behavior for common childhood illnesses such as diarrhea, malaria, and acute respiratory infections. Together, these indicators serve as vital markers of the effectiveness, equity, and accessibility of the health system, particularly for vulnerable populations such as women and children.

Quantitatively, the findings show modest but positive progress across multiple indicators. Most significantly, the under-five mortality rate declined from 132 deaths per 1,000 live births in 2018 to 102 per 1,000 live births in 2023. This decline reflects increasing community awareness, improved child health interventions, and scaled-up immunization efforts in selected regions. Immunization coverage itself improved slightly for DPT3, measles, and pentavalent vaccines, though the gains remain below the World Health Organization’s 90% herd immunity threshold. Exclusive breastfeeding rates increased from 29% to 34%, and stunting declined from 37% to 32%, signaling progress in infant feeding practices and early childhood nutrition. Similarly, the proportion of women attending four or more ANC visits increased from 57% to 68%, while skilled birth attendance rose from 43% to 52%, indicating greater institutional delivery uptake.

However, maternal mortality could not be re-evaluated due to the unavailability of updated national estimates in the 2023–24 NDHS report. This limitation significantly hinders the ability to monitor progress on SDG 3.1 and complicates the evaluation of maternal health programs. Even among indicators that improved, none met international benchmarks or equity-based targets. Immunization, ANC, and skilled birth coverage remain particularly low in Nigeria’s northern, rural, and conflict-affected zones. These findings highlight the persistence of regional disparities and underscore the challenge of ensuring equitable healthcare delivery across diverse geographic and socio-economic settings.

The study’s qualitative analysis offered deeper insights into the systemic and contextual challenges that shape these outcomes. Thematic coding of NDHS narrative sections through NVivo revealed four major themes: geographic and infrastructural access, socioeconomic constraints, community trust and service engagement, and program implementation gaps. Long travel distances, poor road networks, and the lack of functional referral systems were cited as key barriers, particularly in rural areas. Socioeconomic challenges such as poverty, out-of-pocket costs, and food insecurity were consistently mentioned as reasons for delayed or forgone care. Even when services were theoretically “free,” hidden costs such as transportation and unofficial facility fees served as deterrents.

Equally important was the theme of trust and engagement. Communities where health workers were consistent, respectful, and locally known reported higher uptake of services including immunization and ANC. The presence of Ward Health Committees, Health Facility Development Committees, and community health volunteers was shown to improve both health literacy and service utilization. In contrast, where service delivery was sporadic, health workers were unavailable, or communication was poor, families reported lower confidence in formal health services. This reinforces the need to build not just infrastructure, but relationships and trust within communities.

The study also revealed that while Nigeria has developed and launched numerous health policies including the Basic Health Care Provision Fund (BHCPF), the Integrated Maternal, Newborn and Child Health (IMNCH) Strategy, and the National Strategic Health Development Plan (NSHDP) implementation remains inconsistent. The quality and reach of these programs vary widely across states and local government areas, often reflecting differences in subnational leadership, fiscal capacity, and political will. This inconsistency undermines the potential of otherwise well-designed interventions and further entrenches inequities. A key example is the disparity in access to the BHCPF: while some health facilities were fully utilizing funds to support service delivery, others reported delays, underutilization, or complete exclusion from the program.

Data systems and governance emerged as another critical area for improvement. While the NDHS remains the gold standard for national health data, the delay in releasing complete reports and the absence of granular, disaggregated data at the state or local level limit its usefulness for real-time planning. Without timely, actionable data, subnational decision-makers struggle to allocate resources effectively or monitor local performance. Additionally, the lack of maternal mortality data in the 2023–24 preliminary release represents a major gap in Nigeria’s health information system and limits international comparability.

The study also surfaced the need for stronger coordination across the tiers of government. Nigeria’s health system is highly decentralized, with federal, state, and local governments sharing responsibilities. While this structure is intended to bring decision-making closer to the people, it often leads to duplication, fragmentation, and gaps in accountability. Strengthening health governance at all levels particularly in aligning national policies with state-level implementation is essential for improving service delivery and tracking progress toward health goals.

Moreover, the findings highlight that technical fixes alone such as expanding coverage will not be sufficient. Maternal and child health outcomes are deeply influenced by social determinants such as education, gender equity, and household income. Addressing these broader determinants requires cross-sectorial collaboration, including linkages with education, agriculture, and social protection programs. For example, cash transfers and maternity support programs have shown promise in increasing ANC attendance and facility-based deliveries, yet they are not widely scaled or institutionalized across all regions.

Taken together, the study underscores that while Nigeria is making progress, it is not yet on a trajectory that will allow it to meet SDG 3.1 and 3.2 by 2030. The quantitative data show incremental gains, but the qualitative insights make clear that many systemic barriers remain. These include infrastructural deficits, financial barriers, inconsistent service quality, limited community engagement, and inadequate health governance. More importantly, the benefits of progress are not equitably shared, leaving the most vulnerable rural women, low-income households, and conflict-affected populations at the greatest risk of poor health outcomes.

In conclusion, this study presents a mixed and nuanced picture of maternal and child health in Nigeria. It confirms that meaningful progress is possible, as shown by improvements in child mortality, immunization coverage, and service utilization. However, the pace of improvement remains insufficient, and the persistence of regional, socio-economic, and systemic disparities undermines national progress. Going forward, a dual focus is required: first, on strengthening the health system’s core functions workforce, supply chains, financing, data, and infrastructure and second, on building trust, equity, and accountability into every level of service delivery. With targeted investments, policy coherence, and community-centered programming, Nigeria can accelerate its trajectory toward achieving equitable and sustainable maternal and child health outcomes.

## 6.0 RECOMMENDATIONS

To ensure comprehensive and continuous care, antenatal, postnatal, immunization, and nutrition services should be integrated across all levels of maternal and child health delivery. This approach minimizes fragmentation and improves health outcomes.

Frontline health workers should receive regular, targeted training in maternal nutrition, respectful care, and culturally competent service delivery. Improved service quality and communication will enhance trust and patient retention.

Deploying community health workers in underserved areas remains essential for extending care, improving surveillance, and delivering health education. Their proximity to the community boosts early care-seeking behavior.

Digital health solutions such as electronic medical records and mobile health applications should be scaled to improve referrals, performance tracking, and service responsiveness, especially in rural areas.

Government infrastructure investment should focus on high-burden regions with low service coverage. Equitable distribution of resources will help reduce regional disparities in maternal and child health.

The Integrated Maternal, Newborn, and Child Health (IMNCH) strategy should be fully implemented and locally owned, with sustainable funding and active community engagement structures (Mordecai, 2024).

To reduce demand-side barriers, targeted social protection programs such as conditional cash transfers, maternity transport vouchers, and birth kits should be institutionalized nationwide (Mordecai, 2024).

Expanding the National Health Insurance Scheme to fully include maternal and newborn services will reduce out-of-pocket costs and improve access to skilled care for low-income families (Mordecai, 2024).

Federal, state, and local governments must operate within a unified implementation framework to eliminate fragmentation. Clear responsibilities and shared accountability will enhance delivery efficiency.

Timely release of disaggregated health survey data such as NDHS should be enforced. Granular data will improve planning, equity-focused targeting, and local health system responsiveness.

A results-based monitoring and evaluation framework aligned with SDG targets should be adopted. This will enable real-time tracking and informed adjustment of programs (Mordecai, 2024).

## Data Availability

All data produced in the present work are contained in the manuscript

## REFERENCES

Adedoyin, O., Ibrahim, K., & Adebayo, A. (2023). Improving maternal health services in fragile settings: The Nigerian experience. African Journal of Social Health Research, 8(2), 54–66.

Afolabi, S. M., Oladipo, A., & Ekechi, O. (2023). Rural–urban disparities in child survival in Nigeria: Policy implications. AJBSR, 10(4), 32–45.

Agbonle, B. I., Oseji, M. I., Essien, G. D., Swende, L. T., Emma-Nzekwue, N., Idih, E. E., … & Abdulqadir, F. Y. (2022). Improving Maternal, Newborn and Child Health Care in Nigeria–A Medical Women’s Association of Nigeria Multi-Centre Study. Journal of the Medical Women’s Association of Nigeria, 7(2), 55–71.

Ajayi, M. O., Usman, A., & Nwachukwu, B. (2023). Health system performance and maternal mortality in Nigeria: An updated analysis. Nigerian Journal of Public Health, 15(1), 22–34.

Akintunde, T. Y., Salihu, S., & Bello, I. (2023). Trends in skilled birth attendance in Nigeria: A five-year review. F1000Research, 12, 781.

Akpan, E., Chukwuma, E., & Zubair, A. (2024). SDG-aligned maternal health interventions in Nigeria: Gaps and innovations. Journal of Global Development Practice, 14(1), 18–29.

Arksey, H., & O’Malley, L. (2005). Scoping studies: Towards a methodological framework. International Journal of Social Research Methodology, 8(1), 19–32. 10.1080/1364557032000119616

Babajide, O. O., Akinyemi, J. O., & Ayeni, O. (2023). Subnational estimates of maternal mortality in Nigeria: Secondary Data Analysis of female siblings’ survivorship histories. African Journal of Reproductive Health, 27(10), 133–147.

Babatunde, M., Yusuf, T., & Garba, A. (2023). COVID-19 vaccination success and its implications for public health systems in Nigeria. AJBSR, 11(1), 55–68.

Chidinma, G. A. C. (2024). Public Health Expenditure and Maternal Mortality in Nigeria. European Journal of Public Health Studies, 7(1).

Elemuwa, C., Nwosu, I., & Gimba, J. (2024). Community dialogue and maternal health outcomes in Nigeria. Community Dialogue Review, 7(3), 41–58.

Federal Ministry of Health and Social Welfare of Nigeria (FMoHSW), National Population Commission (NPC) [Nigeria], and ICF. (2024). Nigeria Demographic and Health Survey 2023–24: Key Indicators Report. Abuja, Nigeria, and Rockville, Maryland, USA: NPC and ICF.

Gyang, Y., Teryila, K., & Dogo, B. (2023). Health workforce distribution and maternal outcomes in northern Nigeria. Journal of Health Systems & Policy, 9(2), 89–104.

Jeremiah, G. J., Mordecai, O., & Naguibou, D. M. (2023). Identifying the side effects of self-medicated diclofenac sodium among construction workers. Journal of Global Issues and Interdisciplinary Studies, 1(1), 71–75. https://www.researchgate.net/publication/374777423

Joanna Briggs Institute (JBI). (2020). Checklist for Analytical Cross Sectional Studies. The University of Adelaide, JBI.

Kareem, Y. O., Abubakar, Z., Adelekan, B., Ameyaw, E. K., Gbagbo, F. Y., Goldson, E., … & Yaya, S. (2023). Prevalence, trends, and factors associated with teen motherhood in Nigeria: an analysis of the2008–2018 Nigeria demographic and health surveys. International Journal of Sexual Health, 35(2), 248–262.

Kitila, S. B., Feyissa, G. T., Olika, A. K., & Wordofa, M. A. (2022). Maternal healthcare in low-and middle-income countries: a scoping review. Health Services Insights, 15, 11786329221100310.

Levac, D., Colquhoun, H., & O’Brien, K. K. (2010). Scoping studies: Advancing the methodology. Implementation Science, 5(1), 69. 10.1186/1748-5908-5-69

Mordecai, et al., (2024). Analysing Nigeria’s Journey Towards Sustainable Development Goals: A Comprehensive Review From Inception To Present. Article on Health Policy, 10.12688/f1000research.148020.1, 5(30).

Mordecai, O. (2023). A Ten-Year Retrospective Study Evaluating the Effectiveness of the Maternal New-born and Child Health Weeks Intervention among Pregnant Women in Nigeria, https://www.researchgate.net/publication/373949175, 3(1), 17–30.

Mordecai, O. and Jeremiah G. 0 (2023). The Impact of Poverty on Under 5 Children in Rural Communities of the West African Region Alliance for Sustainable Development. https://www.researchgate.net/publication/374255751, 12, 645.

Mordecai, O. et al., (2023). Community Health Management Through Community Dialogue. https://www.researchgate.net/publication/374776657, 7,8,9.

Morufu, R. O., Ajibola, M. A., & Dauda, T. (2023). Environmental health and maternal outcomes in urban slums of Lagos. IJTMRPH, 15(2), 23–36.

National Population Commission (NPC) [Nigeria] and ICF. (2019). Nigeria Demographic and Health Survey 2018. Abuja, Nigeria, and Rockville, Maryland, USA: NPC and ICF.

NPC & ICF. (2019). Nigeria Demographic and Health Survey 2018. Abuja, Nigeria: National Population Commission and ICF International.

Okeke, C., Ezenwaka, U., Ekenna, A., Onyedinma, C., & Onwujekwe, O. (2023). Analysing the progress in service delivery towards achieving universal health coverage in Nigeria: a scoping review. BMC Health Services Research, 23(1), 1094.

Peters, M. D. J., Godfrey, C. M., Khalil, H., McInerney, P., Parker, D., & Soares, C. B. (2015). Guidance for conducting systematic scoping reviews. International Journal of Evidence-Based Healthcare, 13(3), 141–146.

Rotimi, O., Benjamine, O., & Tolulope, M. (2024). Analysis of maternal mortality issues in Nigerian newspapers: a study of South-West, Nigeria (2019–2022). International Journal of African Sustainable Development Research.

Tricco, A. C., Lillie, E., Zarin, W., O’Brien, K. K., Colquhoun, H., Levac, D., … & Straus, S.E. (2018). PRISMA Extension for Scoping Reviews (PRISMA-ScR): Checklist and explanation. Annals of Internal Medicine, 169(7), 467–473.

Umesi, L. U. (2023). Underlying and state-level contextual determinants of early childhood mortality in Nigeria (Doctoral dissertation, The University of Waikato).

Wooldridge, J. M. (2015). Introductory econometrics: A modern approach (5th ed.). Cengage Learning.

